# Improved beta-binomial estimation for reliability of healthcare quality measures

**DOI:** 10.1101/2023.01.07.22283371

**Authors:** Guohai Zhou, Zhenqiu Lin

## Abstract

**Background:** The popular beta-binomial approach to estimate the reliability of healthcare quality measures (Adams et al. 2010 *New England Journal of Medicine*) may yield grossly over-estimated reliabilities for providers with event rates equal to 0% or 100%.

**Objective:** Improve the beta-binomial approach to yield more reasonable reliability estimates for providers with event rates equal to 0% or 100%.

**Method:** We revise the beta-binomial approach by substituting Bayesian estimates with various priors for the crude event rates. We evaluate the new reliability estimates using Monte Carlo studies and two real-world measure examples.

**Results and conclusion:** The revised beta-binomial approach based on Jeffreys non-informative prior yields more reasonable reliability estimates for providers with event rates equal to 0% or 100% and statistically outperforms the original beta-binomial approach regarding bias and standard errors.

## 1 Introduction

The past decade has witnessed the widespread and growing use of healthcare quality measures to evaluate providers’ performance (e.g., hospitals, physicians, physician groups, etc.) in various accountability programs (e.g., the Hospital Readmissions Reduction Program [1]). One fundamental question is how to estimate the reliability of a healthcare quality measure. Reliability quantifies the degree to which the healthcare quality measure reflects providers’ quality of care [2]. Reliability takes a value between zero and one and quantifies the proportion of between-provider differences attributable to the quality of care.

Reliability estimation is of fundamental importance for the usefulness of a healthcare quality measure. For example, when using quality measures for physician-level costs of care for various conditions (e.g., diabetes, heart attack, vascular surgery, etc.),

Adams et al. [3] demonstrated that up to 67% of physicians might be misclassified to a lower cost quartile, and up to 22% of physicians may be misclassified to a higher cost quartile, directly as a consequence of inadequate measure reliability.

One popular approach to estimate the reliability of healthcare quality measures, proposed by Adams [2], was to use a beta-binomial model to estimate the between-provider variance and binomial distributions to estimate the within-provider variance. This beta-binomial approach has been widely used in the literature [3, 5-8, etc.] and is endorsed by the National Quality Forum.

This article aims to revise the beta-binomial approach so providers with event rates equal to 0% or 100% will not get misleadingly optimistic reliability estimates. Section 2 reviews the original beta-binomial approach and illustrates its optimistic bias for providers with event rates equal to 0% or 100%. In Section 3, we propose three revised beta-binomial approaches substituting Bayesian estimates with various priors for crude event rates in the original beta-binomial approach. Sections 4 and 5 will compare the three revised beta-binomial with the original beta-binomial estimate via real data examples and Monte Carlo studies. Section 6 concludes the article and suggests directions for future methodological research.

## 2. Review of the original beta-binomial approach

The original beta-binomial approach first estimates provider-level reliability as follows.

- Fit a beta-binomial model to the provider-level unadjusted event rates data to obtain an estimate for the between-provider variance 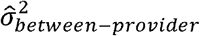.
- For provider i, estimate the within-provider variance by

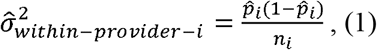

where *n*_*i*_ and 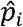 respectively denote the volume and crude event rate.
- For provider i, estimate the reliability by

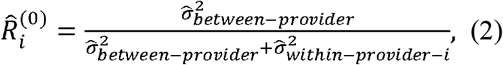

Surprisingly not discussed in previous literature, the original beta-binomial approach suffered from a methodological limitation: providers with event rates 0% or 100% will always have estimated reliability 1, since 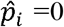 or 1 always yields 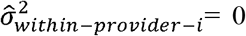 in equation (1) and thus *R*_*i*_=1 in equation (2), even for very small *n*_*i*_. This limitation will make the original beta-binomial approach susceptible to artificially optimistic reliability estimates for small or moderate size providers which happen to have event rates equal to 0% or 100%. In practice, it is not uncommon to see small or moderate size providers with event rates equal to 0% or 100%. For example, providers may happen to have 100% event rate in measures related to severe diseases like cancer; or may happen to have 0% event rate in measures related to rare adverse events. Such providers will get perfect reliability estimates using the original beta-binomial approach. In the next section, we will propose three beta-binomial revised reliability estimates that overcome this limitation so that providers with event rates equal to 0% or 100% will not get artificially perfect reliability estimates.

## 3 Three revised beta-binomial approaches

Let 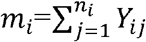 denote the count of events from provider i. We proposed to replace the crude event rate 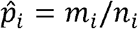 in equation (1) by three Bayesian-based estimators described in Razzaghi [10]. The idea is to use a beta(a,b) prior distribution for *p*_*i*_, provider i’s true measure score, and consider optimal posterior estimates under the squared error loss. Specifically, using the three popular priors as summarized in Razzaghi [10]: 1) uniform non-informative prior beta(1,1), 2) the Jeffreys non-informative prior beta(0.5,0.5) and 3) the informative prior beta 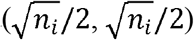 with informativeness coming only from volume, we respectively obtain

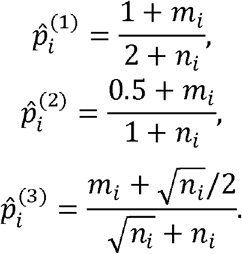

The critical common advantage for 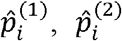 and 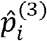over 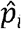 is that when *m*_*i*_=0 (event rate 0%) or *m*_*i*_ = *n*_*i*_ (event rate 100%), they will not automatically produce within-provider variance estimates of 0’s and hence are not susceptible to artificially optimistic reliability estimates. In the next two sections, we will evaluate the following three revised beta-binomial reliability estimates using real-world measure examples and Monte Carlo studies:

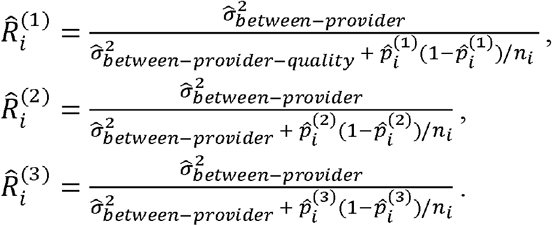

## 4 Real-world measure examples

### 4.1 Example with event rate 0%

We consider the retained surgical item or unretrieved device fragment measure data for 58 California counties from 2005 to 2015 [11], maintained by the state Department of Health Care Access and Information, as part of the broader hospitalization counts and rates of selected adverse hospital events measure. Table 1 presents the raw data from the Lake country. For each year, we applied the original and the three revised beta-binomial approaches to estimate the reliability. Figure 1 visualizes the longitudinal trend of reliability estimates for the Lake county. The original beta-binomial approach suggests unreasonably large longitudinal variation in reliability, namely, from below 0.25 to 1, in adjacent years (such as 2006-2008, and 2012-2014). This is hard to believe given that the underlying factors driving the quality of care (e.g., number of nurses) could not practically change dramatically in a short period for the same measure unit. In addition, the true reliability should not substantially decrease from 1 to 0.003 between 2007 and 2008, when the volume increases from 50130 to 50505. In contrast, all three revised beta-binomial approaches yield reasonable longitudinal variation in the estimated reliability. Furthermore, they all agree very well with the original beta-binomial approach at years other than 2007 and 2013 (when the original beta-binomial approach incorrectly yields reliability estimate 1 due to the event rate equal to 0%).

**Table 1.** Retained surgical item or unretrieved device fragment_raw measure data from the Lake county 2005 - 2015.

| Year | Number of cases | Number of events | Crude event rate per 100K (measure score) |
| --- | --- | --- | --- |
| 2005 | 4 | 49529 | 8.08 |
| 2006 | 4 | 49985 | 8 |
| 2007 | 0 | 50130 | 0 |
| 2008 | 2 | 50505 | 3.96 |
| 2009 | 4 | 50721 | 7.89 |
| 2010 | 1 | 51151 | 1.95 |
| 2011 | 1 | 51085 | 1.96 |
| 2012 | 1 | 51501 | 1.94 |
| 2013 | 0 | 51581 | 0 |
| 2014 | 3 | 52953 | 5.67 |
| 2015 | 1 | 39003 | 2.56 |

**Figure 1:**
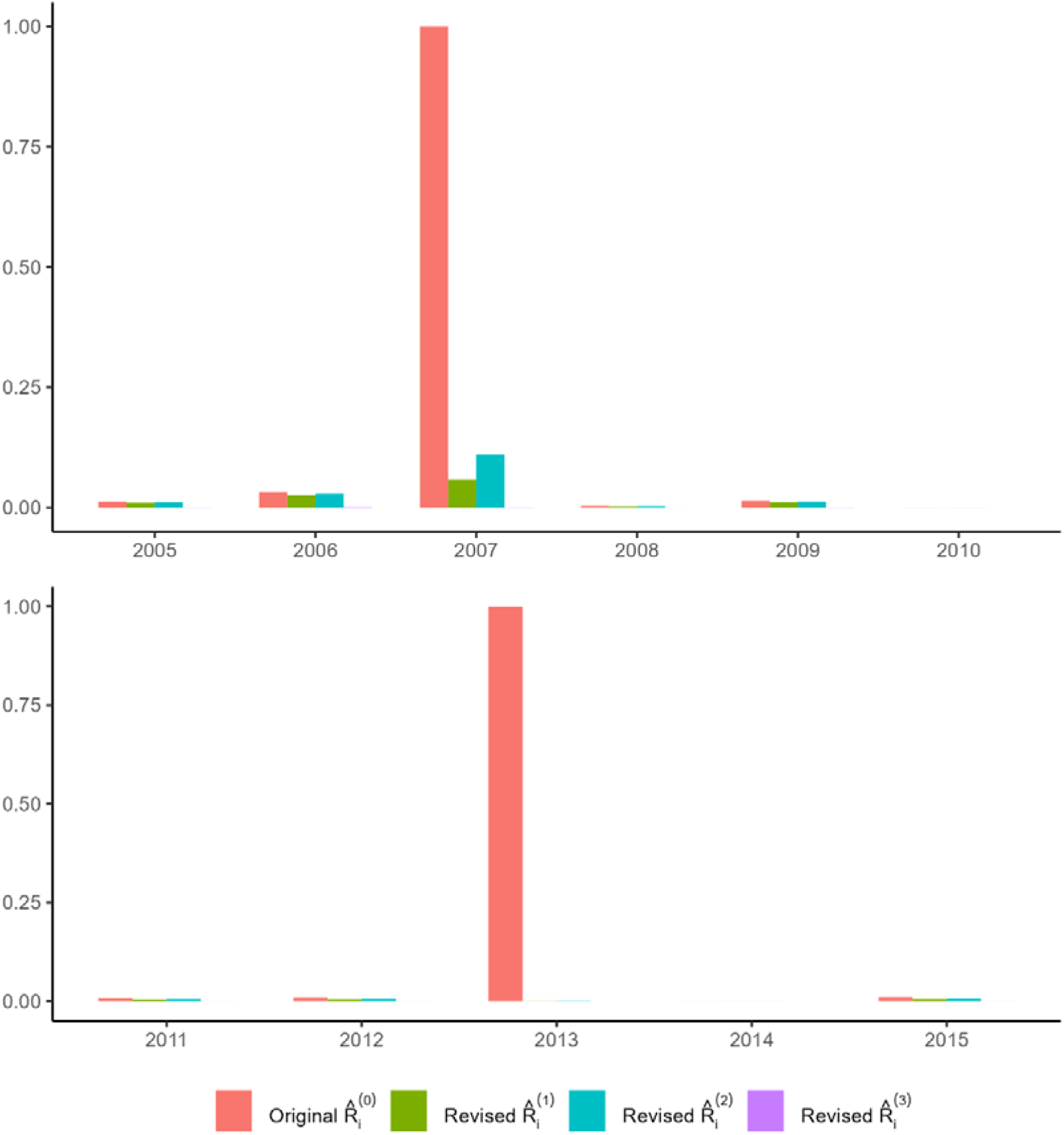
Reliability estimates (y-axis) based on the original and the three revised beta-binomial approaches for the Lake county.

### 4.2 Example with event rate 100%

We also consider the adjuvant chemotherapy colon cancer (ACCC) measure for prospective payment system exempt cancer hospitals (data downloaded from Hospital Compare in 2018 [12]). Table 2 presents the raw provider-level data.

**Table 2:**
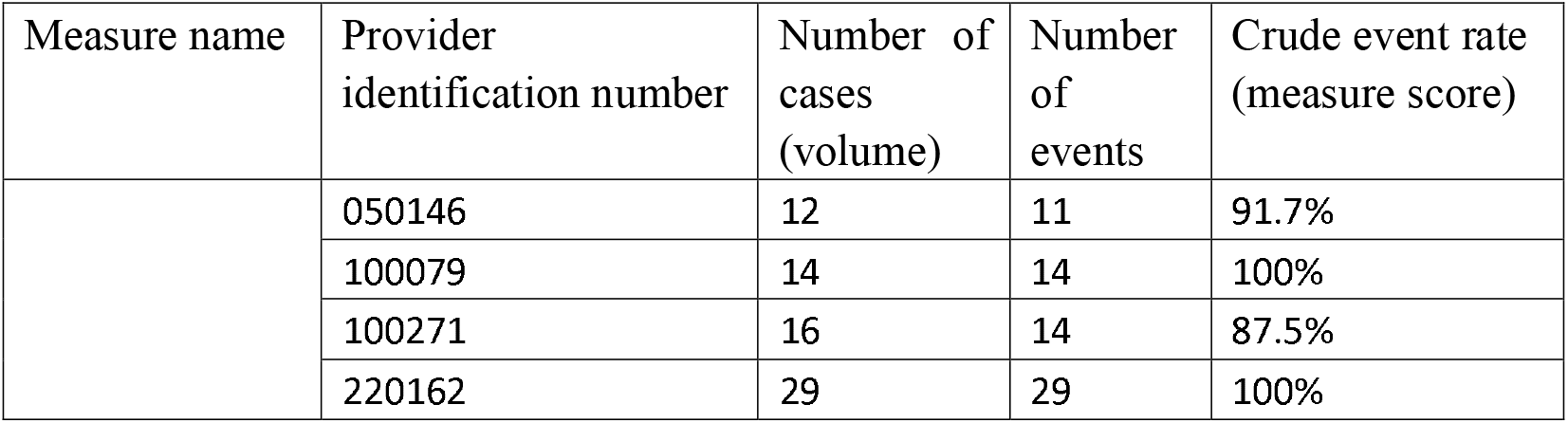
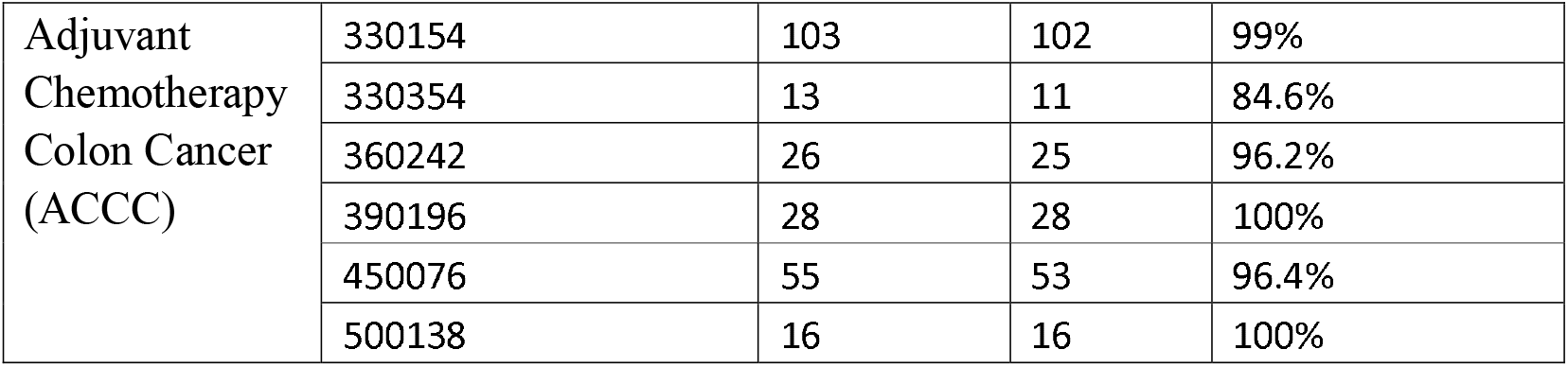
Adjuvant chemotherapy colon cancer measure score for year 2017 for PPS exempt Cancer hospital

We calculate reliability using the original and the three revised beta-binomial approaches and present their comparisons in Figure 2, with the categories for crude event rate and volume chosen based on the distribution of real data in Table 2. Figure 2 indicates that for the four providers with 100% crude event rates, the original beta-binomial approach yields misleadingly perfect reliability estimates (=1’s), whereas the revised beta-binomial approach 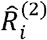 yields more reasonable reliability estimates with an as-expected property of monotonically increasing with volume. For other providers, the revised beta-binomial approach 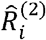 agrees very well with the original beta-binomial approach.

**Figure 2:**
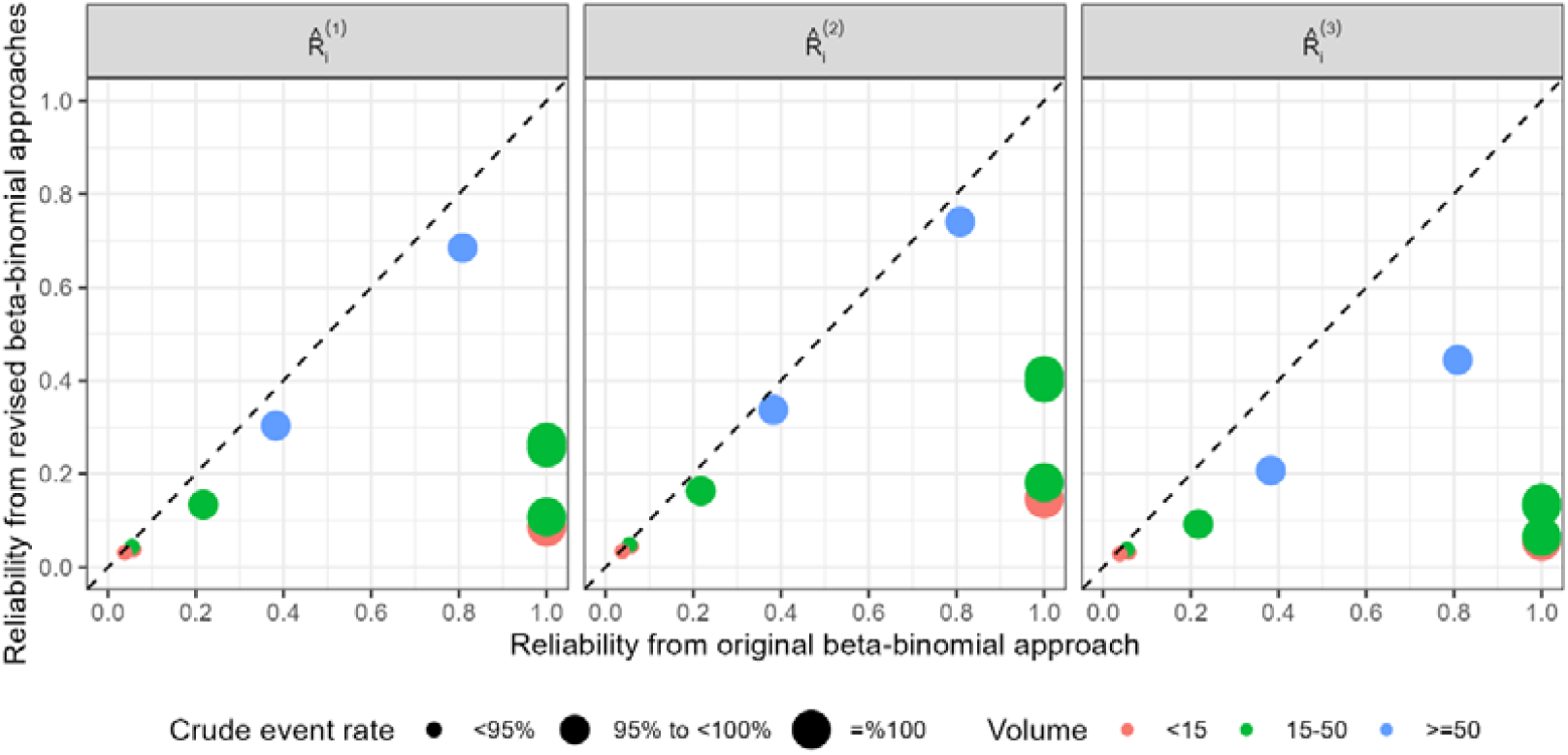
Reliability estimates from the original and revised beta-binomial approaches (line of identity dashed).

Overall, the results from the two above real data examples appear to both suggest that the revised beta-binomial approach 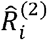 outperforms the original beta-binomial approach 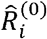 because it produces more reasonable reliability estimates for providers with an event rate of 0% or 100%, for which the original beta-binomial always over-estimate the reliability to 1. We will compare different reliability estimation approaches and confirm the advantage of the revised beta-binomial approach 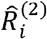 using Monte Carlo studies in the next section.

## 5 Empirical comparisons: Monte Carlo studies

We conduct Monte Carlo studies, in which the true provider-level reliabilities are known, to compare the original and three revised beta-binomial approaches in terms of the bias and standard errors (SEs).

Specifically, we independently simulate true measure scores from a beta distribution with parameters 77.8 and 2.68 for each provider. This beta distribution is estimated based on the real data from the ACCC measure. The true between-provider variance is then known to be 0.000394 based on this beta distribution. For a provider with volume n and true measure score p, the true within-provider variance is then known to be p(1-p)/n. Therefore, the true provider-level reliability for this provider is known to be 0.000394/(0.000394+p(1-p)/n).

We consider three scenarios for the numbers of providers in the measure cohort: n=10 (the number of providers in the ACCC measure), 100 and 1000. We reused each provider in the ACCC measure 10 and 100 times respectively for the latter two. We use the real volume from the ACCC measure, simulate the number of events based on binomial probabilities, and stratify results by categories of crude event rate. We also consider two scenarios for provider volume: equal to the real volume from ACCC measure, or multiply the real volume from the ACCC measure by 10.

Table 3 displays the results of empirical bias and SEs associated with the estimation for provider-level reliability. The original beta-binomial approach yields large bias and zero SEs for the provider-level reliability estimation for providers with crude event rate equal to 100%, regardless of how large the volume or the number of providers is. Therefore, the original beta-binomial approach’s susceptibility to artificially optimistic reliability estimates for providers with event rates equal to 0% or 100% is indeed statistically inappropriate. In contrast, all three revised beta-binomial approaches improve this susceptibility, and the revised beta-binomial approach based on 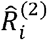 appears to deliver the lowest bias. For providers whose crude event rates are not 100%, the revised beta-binomial approach based on 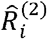 roughly matches the original beta-binomial approach’s performance in terms of bias and SEs, which further supports its potential to replace the original beta-binomial approach.

**Table 3:** **Empirical bias and standard errors (SEs) associated with provider-level reliability estimates using different approaches, based on 1000 simulation iterations. True measure scores generated from a beta distribution with parameter values set to estimates based on the ACCC measure real data**

| Total number of providers | Crude event rate category | Average number (%)* of providers in each crude rate category | Bias (SE) associated with *provider-level* reliability estimates using different approaches |  |  |  |
| --- | --- | --- | --- | --- | --- | --- |
|  |  |  | Original beta-bino mial approach | Revised beta-binomial approach based on |  |  |
| | | | | $\hat{R}_i^{(1)}$ | $\hat{R}_i^{(2)}$ | $\hat{R}_i^{(3)}$ |
| Provider volume equal to real volume from ACCC Measure |  |  |  |  |  |  |
| 10 | =100% | 4.9 (48.9%) | 0.72 (0.00) | -0.15 (0.20) | -0.11 (0.25) | -0.20 (0.15) |
|  | 95%-100% | 2.3 (22.8%) | -0.25 (0.28) | -0.28 (0.20) | -0.27 (0.26) | -0.32 (0.19) |
|  | <95% | 3.0 (30.3%) | -0.11 (0.15) | -0.12 (0.20) | -0.12 (0.15) | -0.13 (0.12) |
| 100 | = 100% | 49.0 (49.0%) | 0.72 (0.00) | -0.12 (0.17) | -0.05 (0.20) | -0.19 (0.09) |
|  | 95%-100% | 21.2 (21.2%) | -0.06 (0.22) | -0.14 (0.17) | -0.11 (0.22) | -0.23 (0.15) |
|  | <95% | 29.8 (29.8%) | -0.09 (0.11) | -0.11 (0.17) | -0.10 (0.10) | -0.13 (0.08) |
| 1000 | =100% | 490.2 (49.0%) | 0.72 (0.00) | -0.11 (0.17) | -0.03 (0.18) | -0.19 (0.08) |
|  | 95%-100% | 212.6 (21.3%) | -0.01 (0.19) | -0.11 (0.17) | -0.07 (0.19) | -0.21 (0.13) |
|  | <95% | 297.2 (29.7%) | -0.08 (0.09) | -0.11 (0.17) | -0.10 (0.09) | -0.13 (0.07) |
| Provider volume equal to real volume from ACCC Measure *multiplied by 10* |  |  |  |  |  |  |
| 10 | =100% | 1.2 (11.7%) | 0.16 (0.00) | -0.01 (0.26) | 0.05 (0.18) | -0.27 (0.22) |
|  | 95%-100% | 7.7 (76.8%) | -0.13 (0.27) | -0.16 (0.26) | -0.15 (0.27) | -0.27 (0.26) |
|  | <95% | 2.2 (22.0%) | -0.06 (0.22) | -0.07 (0.26) | -0.07 (0.22) | -0.13 (0.23) |
| 100 | =100% | 4.0 (4.0%) | 0.15 (0.00) | 0.05 (0.16) | 0.10 (0.03) | -0.23 (0.11) |
|  | 95%-100% | 77.1 (77.1%) | 0.00 (0.13) | -0.03 (0.16) | -0.01 (0.14) | -0.15 (0.17) |
|  | <95% | 19.0 (19.0%) | -0.04 (0.16) | -0.05 (0.16) | -0.05 (0.16) | -0.11 (0.18) |
| 1000 | =100% | 38.6 (3.9%) | 0.16 (0.00) | 0.05 (0.16) | 0.10 (0.02) | -0.22 (0.09) |
|  | 95%-100% | 771.9 (77.2%) | 0.01 (0.12) | -0.02 (0.16) | -0.00 (0.13) | -0.14 (0.16) |
|  | <95% | 189.6 (19.0%) | -0.04 (0.15) | -0.05 (0.16) | -0.04 (0.16) | -0.11 (0.17) |

\*Average over the 1000 simulation iterations.

## 6 Discussion

We proposed three revised beta-binomial approaches to estimate healthcare measure reliability. Out of the three, we recommend the revised beta-binomial approach using Bayesian estimates with Jeffreys non-informative prior based on three folds of evidence: 1) it avoids the artificially perfect reliability estimation associated with the original beta-binomial approach for providers with event rates equal to 0% or 100%, 2) it produces more reasonable reliability estimates for providers with event rate equal to 0% or 100% in real world measure examples, and 3) it outperforms the original beta-binomial approach in Monte Carlo studies in terms of bias and standard errors.

This revised beta-binomial approach can be readily adapted to improve the composite quality measure developed in Dimick et al. [13]. Their composite measures used each hospital’s estimated reliability as the weight to compute a weighted average of each hospital’s crude and volume-predicted mortality. They used the same way to calculate within-provider variance for their current reliability calculation as in the original beta-binomial approach. Therefore, their composite measures would be sub-optimal for providers with crude mortality rates equal to 0% or 100%. Based on the revised beta-binomial reliability estimates using Jeffreys non-informative prior, we are working on a formal extension of the composite quality measure developed in Dimick et al. [13].

## Data Availability

All data produced in the present work are contained in the manuscript

## Appendix

The R and SAS code to reproduce all figures and tables can be found at github.com/GuohaiZhou/measure_reliability

